# Hospitalization for self-harm during the early months of the Covid-19 pandemic in France: a nationwide study

**DOI:** 10.1101/2020.12.18.20248480

**Authors:** Fabrice Jollant, Adrien Roussot, Emmanuelle Corruble, Jean-Christophe Chauvet-Gelinier, Bruno Falissard, Yann Mikaeloff, Catherine Quantin

**Author notes:** Correspondence to: Dr. Fabrice Jollant, Hôpital Sainte-Anne, Clinique des Maladies Mentales et de l’Encéphale (CMME), 100 rue de la Santé 75674 Paris cedex 14, France. These two authors contributed equally to this work (Fabrice Jollant, Adrien Roussot).

## Abstract

**Introduction:** Little is known to date about the impact of Covid-19 pandemic on self-harm incidence.

**Methods:** The number of hospitalizations for self-harm in France (mainland and overseas) from January to August 2020 (which includes the first confinement from March 17^th^ to May 11^th^) was compared to the same period in 2019-2017. Hospital data with the ICD-10 codes X60-84 were extracted from the national administrative database (PMSI).

**Results:** There were 53,583 hospitalizations for self-harm in France between January and August 2020. Compared to the same period in 2019, this represents an overall 8.5% decrease. This decrease started the first week of the confinement and the number of hospitalizations remained at lower levels relative to 2019 until the end of August. The decrease was more marked in women (−9.8%) than men (−6.4%). However, an increase in hospitalizations was observed in individuals aged 75 and older (+5.3 to +11.6%). Moreover, the number of self-harm by firearm (+20.3%), jumping from height (+10.5%), and drowning (+4.7%) increased between 2019 and 2020, as well as the number of hospitalizations in intensive care (+3.5%) and deaths at discharge from hospital (+8.0%). No correlation was found between the evolution in the number of hospitalizations for self-harm and the number of severe cases of Covid-19 (hospitalization and mortality rates) across administrative departments.

**Discussion:** During the early months of the Covid-19 pandemic in France - including the first confinement -, a general decrease in the number of hospitalizations for self-harm was observed. However, an increase was found among elderly, a population at higher Covid 19-related mortality risk, and in the number of more severe suicidal acts. These results, therefore, shed light on a complex relationship between the pandemic and self-harm occurrence. This situation may change with time, which requires active suicide prevention strategies.

## INTRODUCTION

What will be later called Covid-19 was first reported as atypical cases of pneumonia in Wuhan, China, at the end of 2019 and was followed by a major outbreak in this province. The causal germ was soon identified as a novel strain of coronavirus named SARS-Cov-2. The disease rapidly spread across the world and was declared a pandemic by the World Health Organization in March 2020. The pandemic is still ongoing at the time of writing this manuscript in December 2020 and has killed more than 1.6 million people worldwide to date. In order to limit the contagion, governments implemented various local and national strategies, the most striking of them being physical distancing and confinement of populations at the scale of a city, a region/state/province, or a whole country. While pandemics and confinement of populations for epidemic reasons is not new in History, it was a totally novel experience for living generations in many countries including France.

Studies of previous quarantines in recent years have reported a negative impact on mental health overall ^1^. Studies conducted during the present Covid-19 pandemic confirmed these findings and showed increased rates of anxiety, depression and traumatic stress across the general population in various countries ^2^. Some studies also showed increases in suicidal ideation ^3, 4^, notably in conditions of lockdown ^5^. In France, a regular web-based survey of a representative sample of the general population conducted by *Santé Publique France* (“Public Heath France”) since March 16th, 2020 (https://www.santepubliquefrance.fr/etudes-et-enquetes/covid-19-une-enquete-pour-suivre-l-evolution-des-comportements-et-de-la-sante-mentale-pendant-l-epidemie) showed an initial increase in depression and anxiety levels associated with sleep problems and alcohol and tobacco use, followed by a relative improvement of these indexes after the end of the lockdown. Finally, concerns have notably been raised concerning specific populations such as adolescents and young adults, unemployed and economically disadvantaged, individuals with a pre-existing mental illness, and frontline medical workers ^4, 6–10^.

Regarding the worrying deterioration of mental health during confinements in the general population - combined with issues pertaining to potentially reduced access to mental health care, limited social support, and for many individuals professional and financial problems - one major question is to know if Covid-19 and related preventative measures such as confinement may have had an effect on the rates of suicidal acts ^11–13^. Few data on the effect of previous epidemics on suicidal behavior exist and studies were usually of low methodological quality ^14, 15^. Preliminary results in the context of the Covid-19 pandemics suggest no significant negative impact in the short-term. Data on suicide deaths are still scarce due to the usual delay in causes-of-mortality data access ^16^. However, countries with real-time collection of suicide data showed decreased or unchanged suicide rates, e.g. in Peru ^17^, Australia ^18^, Norway ^19^, Japan (early period) ^20^, or Massachusetts, USA (Faust et al. in press). Data on self-harm are still limited. In one University hospital in Ireland, presentation for self-harm dropped in March-April before rising in April-May ^21^. In a trauma center in the UK, an increase in self-harm presentation was observed ^22^. Of note, an increase in more violent suicidal acts (penetrative lesions) has also been locally observed ^23^, which incites to analyze the evolution of self-harm occurrence as a function of the suicidal means and lethality.

In France, the first Covid-19 cases were officially detected and publicly announced on January 24^th^, 2020. Initial interventions focused on the isolation of confirmed or suspected cases, and contact cases. The rapidly increasing number of contaminated cases and deaths led the government to several graded decisions during the first half of March (e.g. closing of schools, bars and restaurants) until the implementation of a national lockdown starting on March, 17^th^ (week 11). The whole population was then confined at home and only restricted professionals were allowed to go to work. Schools and universities were closed. The lockdown ended on May 11^th^ (week 19). Importantly, while the lockdown was national, the level of contamination and deaths varied importantly between administrative regions and departments. Deaths and hospitalizations due to Covid-19 remained low during the summer vacation time in July and August although signs of an active viral circulation were reported during this period. A second confinement was then declared starting October 30^th^ until December 15^th^, 2020 in a very different context of economic crisis, awareness of the prolonged presence of the pandemics, uncertainties about its ending, all mixed with expectations of a vaccine.

The present article will focus on the early months of the pandemics in France - therefore comprising the first confinement and the first deconfinement period (including the summer period) - between January and August 2020. We firstly aimed at investigating changes in hospitalization rates for self-harm in comparison to similar periods in 2019 (and additional comparisons to 2018 and 2017). Second, we also focused on more severe acts, i.e. those necessitating intensive care and those using a violent means. These acts have been associated with an increased risk of subsequent suicide ^24^. Finally, we were interested in studying a potential link between local changes in rates of self-harm and the level of contamination in administrative departments of France. The number of positive tests being not a relevant index in a period when testing was not widespread, we measured the number of hospitalizations and deaths related to Covid-19 infection. We hypothesized that the local reality of the pandemics had no influence on self-harm rates.

An important strength of this study is the exhaustive information about hospitalization for severe self-harm (e.g. necessitating a subsequent hospitalization) at a nationwide level.

## METHODS

### Study design and participants

We used the national *Programme de Médicalisation des Systèmes d’Information* (PMSI) database, which collects discharge abstracts for all patients admitted to public and private hospitals in France. In 2020, this database contains the information of approximately 1,700 public hospitals and private clinics.

All hospital stays for self-harm registered in France (including overseas territories) were included in the analysis. The number of monthly hospitalizations in Medicine/Surgery/Obstetrics (MCO) with an associated diagnosis of self-harm (International Classification of Diseases, 10th revision (ICD-10) codes X60 to X84) observed in France between January and August 2020 for patients aged 10 years and more were extracted. We also extracted the hospital stays for self-harm from the same months in 2017, 2018 and 2019. Weekly number of hospitalizations for self-harm were additionally extracted for 2019 and 2020.

Self-harm was furthermore classified as violent (X66 to 82) and non-violent (X60-65) acts as previously done ^24^.

We additionally retrieved information on gender, age, stays in an intensive care unit and hospital death at discharge. Departments of residence were identified according to the zip-code registered for each patient in PMSI data.

We identified all hospital stays for COVID-19 infection registered between January and August 2020 in France with ICD-10 codes U0710, U0711, U0712, U0714 or U0715 as primary diagnoses.

### Statistical and spatial analyses

We reported the number of hospital stays by gender, age group and month at discharge, and calculated evolution in terms of number and rates between 2020 and 2019, 2018, and 2017.

We mapped three indicators at the scale of the French administrative departments:

– the 2019-2020 evolution of hospitalizations for self-harm;
– the standardized hospitalization rates for COVID-19 per 10,000 inhabitants;
– the standardized in-hospital mortality rates for COVID-19 per 10,000 inhabitants.

Hospitalization and in-hospital mortality rates for COVID-19 were standardized for age and gender according to the direct method, using the 2017 national census data produced by The National Institute of Statistics and Economic Studies (INSEE) as reference.

The potential spatial overlap between the department’s distributions of those three variables was analysed using a Pearson correlation test.

All analyses were performed using SAS (SAS Institute Inc, Version 9.4, Cary, NC).

### Ethics approval

The access to the ATIH hospital data platform was approved by the National Committee for data protection (registration number 2204633) and therefore was conducted in accordance with the Declaration of Helsinki. Written consent was not needed for this study.

## RESULTS

A total of 53,584 hospitalizations for self-harm occurred between January and August 2020. This is less than in 2019 (−4,972 representing −8.5%), 2018 (−7,948; −12.9%), and 2017 (−6,528; −10.9%) for the same period (**Table 1**). At the monthly level, this decrease started in March after a relative increase in January and February (except when compared to January 2018). At the weekly level, **Figure 1** shows that the decrease in hospitalization for self-harm in 2020 started abruptly at week 11, the week when the general confinement was declared. The lower level of hospitalizations for self-harm in 2020 persisted from March until August 2020 in spite of an increase between week 13 (therefore during confinement) until approximately week 27 (early July).

**Table 1.** Total number of monthly hospitalizations for self-harm in France in 2020 as compared to 2019, 2018 and 2017 (January to August only).

|  | **2020** | **2019** |  |  | **2018** |  |  | **2017** |  |  |
| --- | --- | --- | --- | --- | --- | --- | --- | --- | --- | --- |
|  | **N** | **N** | **Diff. 2020-2019** | **% change** | **N** | **Diff. 2020-2018** | **% change** | **N** | **Diff. 2020-2017** | **% change** |
| January | 7720 | 7251 | 469 | 6.5 | 7947 | -227 | -2.9 | 7296 | 424 | 5.8 |
| February | 6997 | 6280 | 717 | 11.4 | 6883 | 114 | 1.7 | 6802 | 195 | 2.9 |
| March | 6558 | 7551 | -993 | -13.2 | 7781 | -1223 | -15.7 | 8147 | -1589 | -19.5 |
| April | 5844 | 7406 | -1562 | -21.1 | 7557 | -1713 | -22.7 | 7170 | -1326 | -18.5 |
| May | 6407 | 7936 | -1529 | -19.3 | 8133 | -1726 | -21.2 | 8132 | -1725 | -21.2 |
| June | 6890 | 7581 | -691 | -9.1 | 8234 | -1344 | -16.3 | 7951 | -1061 | -13.3 |
| July | 6914 | 7495 | -581 | -7.8 | 7839 | -925 | -11.8 | 7401 | -487 | -6.6 |
| August | 6254 | 7056 | -802 | -11.4 | 7158 | -904 | -12.6 | 7213 | -959 | -13.3 |
| Total | 53584 | 58556 | -4972 | -8.5 | 61532 | -7948 | -12.9 | 60112 | -6528 | -10.9 |

Footnotes: The confinement lasted from March 17<sup>th</sup> to May 11<sup>th</sup>.

**Figure 1.**
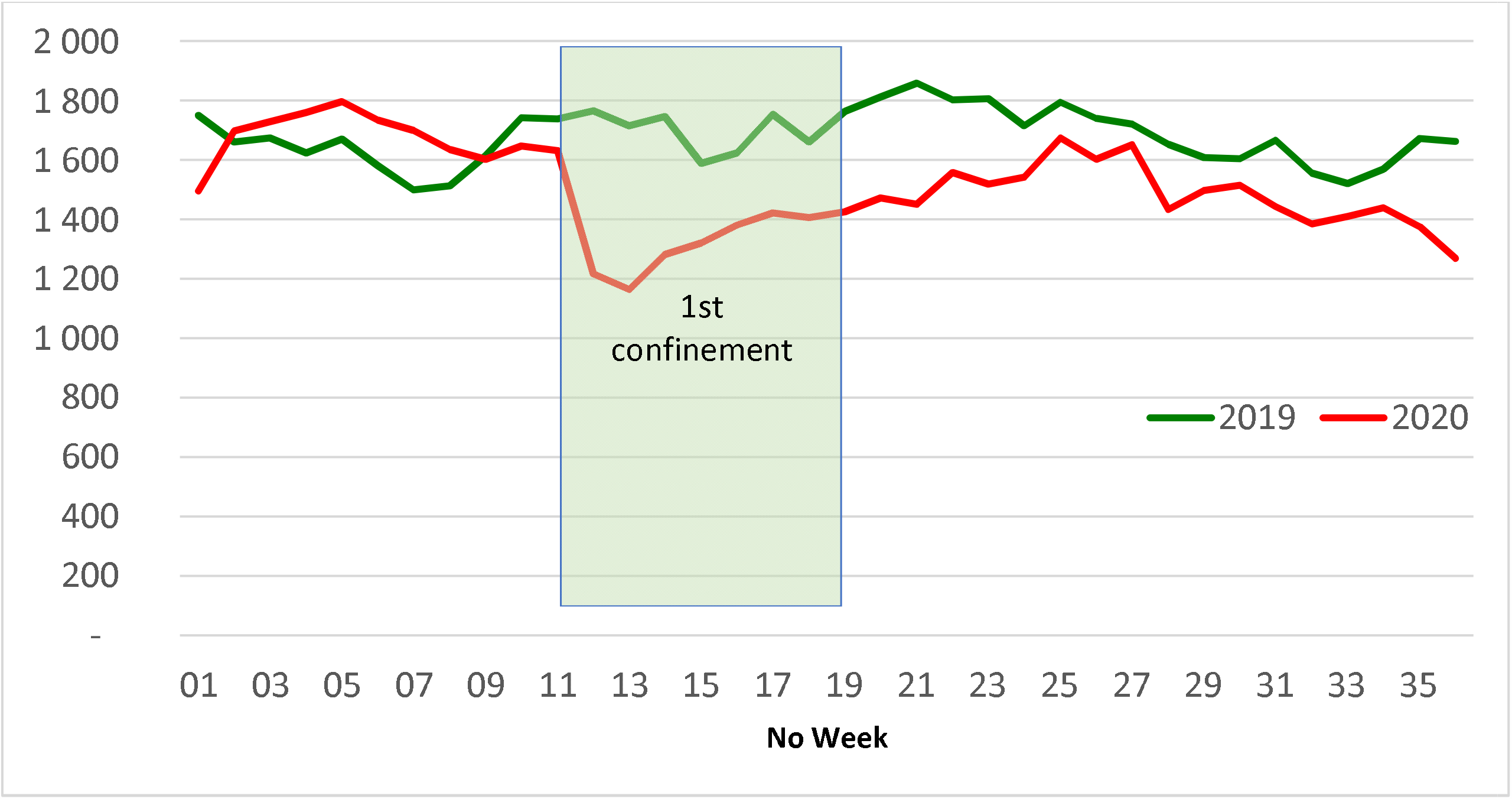
Number of weekly hospitalizations for self-harm in France in 2020 and 2019 (January to August only)

When comparing 2020 to 2019, this decrease was found in both gender, but was more important among women (−9.8%) than men (−6.4%) (**Table 2 and Figure 2**).

**Table 2.** Number of hospitalizations for self-harm in France in 2019 and 2020 (January to August only) per gender and age.

| Age group (year) | N in 2020 |  |  | N in 2019 |  |  | Difference in N (2020-2019) |  |  | % change (2020-2019) |  |  |
| --- | --- | --- | --- | --- | --- | --- | --- | --- | --- | --- | --- | --- |
|  | Men | Women | Total | Men | Women | Total | Men | Women | Total | Men | Women | Total |
| 10-14 | 450 | 2273 | 2723 | 571 | 2924 | 3495 | -121 | -651 | -772 | -21.2 | -22.3 | -22.1 |
| 15-19 | 1632 | 5053 | 6685 | 1943 | 5904 | 7847 | -311 | -851 | -1162 | -16.0 | -14.4 | -14.8 |
| 20-24 | 1774 | 2827 | 4601 | 1883 | 2961 | 4844 | -109 | -134 | -243 | -5.8 | -4.5 | -5.0 |
| 25-29 | 1819 | 2031 | 3850 | 1915 | 2199 | 4114 | -96 | -168 | -264 | -5.0 | -7.6 | -6.4 |
| 30-34 | 2081 | 1922 | 4003 | 2119 | 2187 | 4306 | -38 | -265 | -303 | -1.8 | -12.1 | -7.0 |
| 35-39 | 2196 | 2099 | 4295 | 2333 | 2292 | 4625 | -137 | -193 | -330 | -5.9 | -8.4 | -7.1 |
| 40-44 | 2093 | 2073 | 4166 | 2205 | 2550 | 4755 | -112 | -477 | -589 | -5.1 | -18.7 | -12.4 |
| 45-49 | 2374 | 3037 | 5411 | 2644 | 3426 | 6070 | -270 | -389 | -659 | -10.2 | -11.4 | -10.9 |
| 50-54 | 2040 | 2895 | 4935 | 2199 | 3202 | 5401 | -159 | -307 | -466 | -7.2 | -9.6 | -8.6 |
| 55-59 | 1543 | 2238 | 3781 | 1672 | 2308 | 3980 | -129 | -70 | -199 | -7.7 | -3.0 | -5.0 |
| 60-64 | 971 | 1571 | 2542 | 1063 | 1668 | 2731 | -92 | -97 | -189 | -8.7 | -5.8 | -6.9 |
| 65-69 | 752 | 1229 | 1981 | 723 | 1235 | 1958 | 29 | -6 | 23 | 4.0 | -0.5 | 1.2 |
| 70-74 | 499 | 892 | 1391 | 510 | 908 | 1418 | -11 | -16 | -27 | -2.2 | -1.8 | -1.9 |
| 75-79 | 382 | 607 | 989 | 353 | 586 | 939 | 29 | 21 | 50 | 8.2 | 3.6 | 5.3 |
| 80-84 | 351 | 599 | 950 | 301 | 550 | 851 | 50 | 49 | 99 | 16.6 | 8.9 | 11.6 |
| 85+ | 502 | 779 | 1281 | 504 | 718 | 1222 | -2 | 61 | 59 | -0.4 | 8.5 | 4.8 |
| Total | 21459 | 32125 | 53584 | 22938 | 35618 | 58556 | -1479 | -3493 | -4972 | -6.4 | -9.8 | -8.5 |

**Figure 2.**
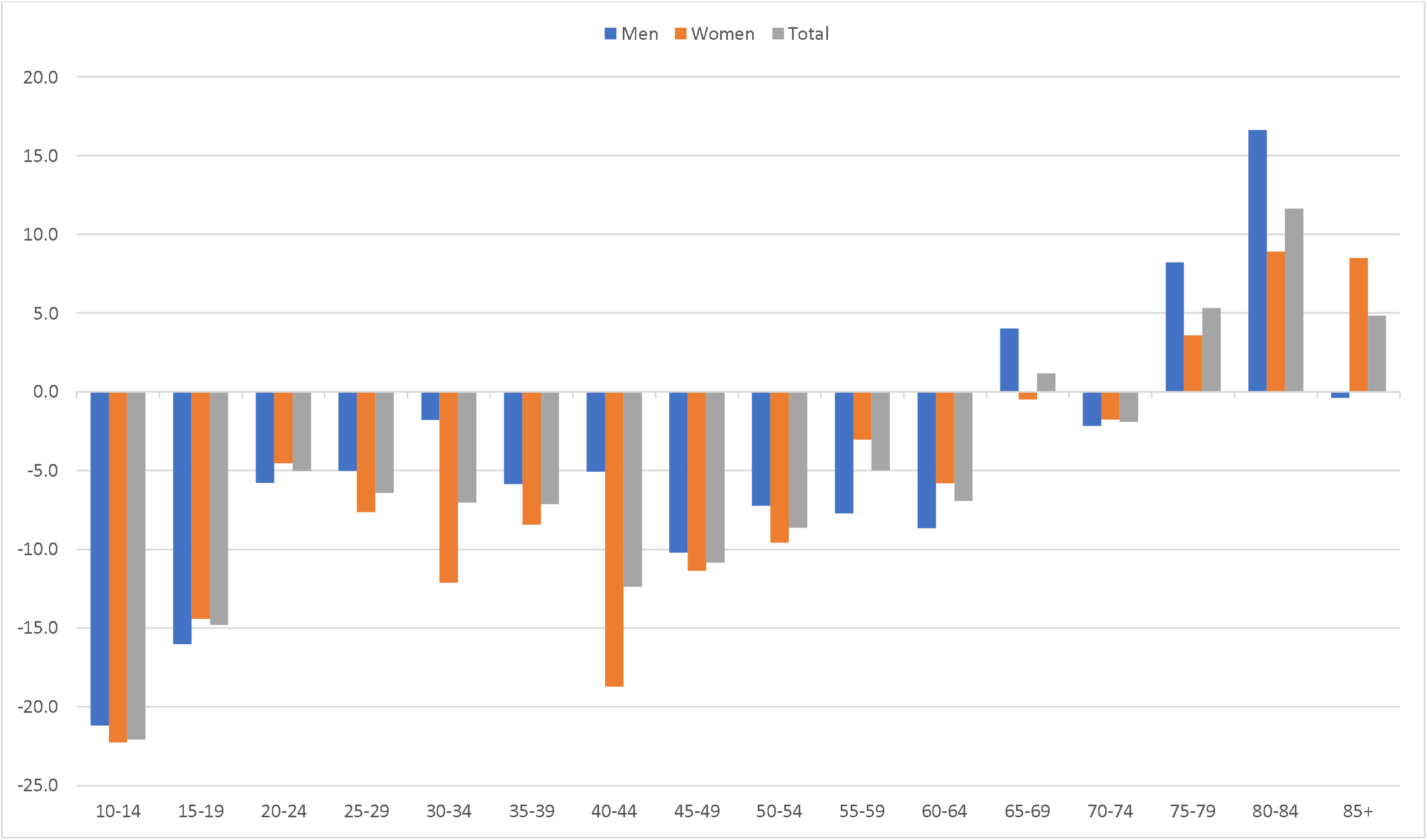
Change (%) in number of hospitalizations for self-harm in France in 2020 as compared to 2019 (January to August only) per age group and gender.

Regarding age, the decrease was found in all age group at the exception of elderly in whom an increase in hospitalizations was found, notably in individuals above 75 years old (+5.3% in 75-79; +11.6% in 80-84; +4.8 in 85+) in both men and women (**Table 2 and Figure 2**).

Decrease in self-harm between 2019 and 2020 was found for all suicidal means at the notable exception of firearm (+20.3%), jumping from height (+10.5%), and drowning (+4.7%) (**Table 3**). Moreover, hospitalization in intensive care for self-harm increased between 2019 and 2020 (+3.5%) as well as the number of deaths at hospital following a self-harm (+8.0%).

**Table 3.** Number of hospitalizations for self-harm in France in 2019 and 2020 (January to August only) according to the means.

| **Means** | **2020** | **2019** | **Difference in N**<br>**between 2020-2019** | **% change**<br>**(2019-2020)** |
| --- | --- | --- | --- | --- |
| Drugs (X60-X64) | 39082 | 42972 | -3890 | -9.1 |
| Alcohol (X65) | 2693 | 2680 | 13 | 0.5 |
| Violent means (X66-X82), including: | 10 012 | 10 571 | -559 | -5.3 |
| *Solvents, carbon monoxide, pesticides, other*<br>*chemicals (X66-X69)* | 1359 | 1449 | -90 | -6.2 |
| *Hanging, strangulation (X70)* | 1658 | 1692 | -34 | -2.0 |
| *Drowning (X71)* | 135 | 129 | 6 | 4.7 |
| *Firearm (X72-X74)* | 356 | 296 | 60 | 20.3 |
| *Fire, explosive (X75-X77)* | 145 | 198 | -53 | -26.8 |
| *Sharp or blunt object (X78-X79)* | 5195 | 5708 | -513 | -9.0 |
| *Jumping from height (X80)* | 1034 | 936 | 98 | 10.5 |
| *Jumping/crashing (X81-X82)* | 130 | 163 | -33 | -20.2 |
| *Other means (X83-X84)* | 1797 | 2333 | -536 | -23.0 |
| Hospitalization in intensive care | 5 894 | 5 693 | 201 | 3.5 |
| Death at discharge | 759 | 703 | 56 | 8.0 |

The North-Eastern quarter of France and the oversea department of French Guyana were more affected by COVID-19 epidemic than other departments between January and August 2020 both in terms of hospitalizations or in-hospital mortality. There was no significant correlation between change in the number of hospitalizations for self-harm and both hospitalizations for Covid-19 (r=-0.1; p=0.3) or in-hospital mortality by Covid-19 (r=-0.1; p=0.2) across administrative departments (**Figure 3**). As expected, hospitalization and in-hospital mortality for COVID-19 rates were strongly intercorrelated (r= 0.9; p< 0.0001).

**Figure 3.**
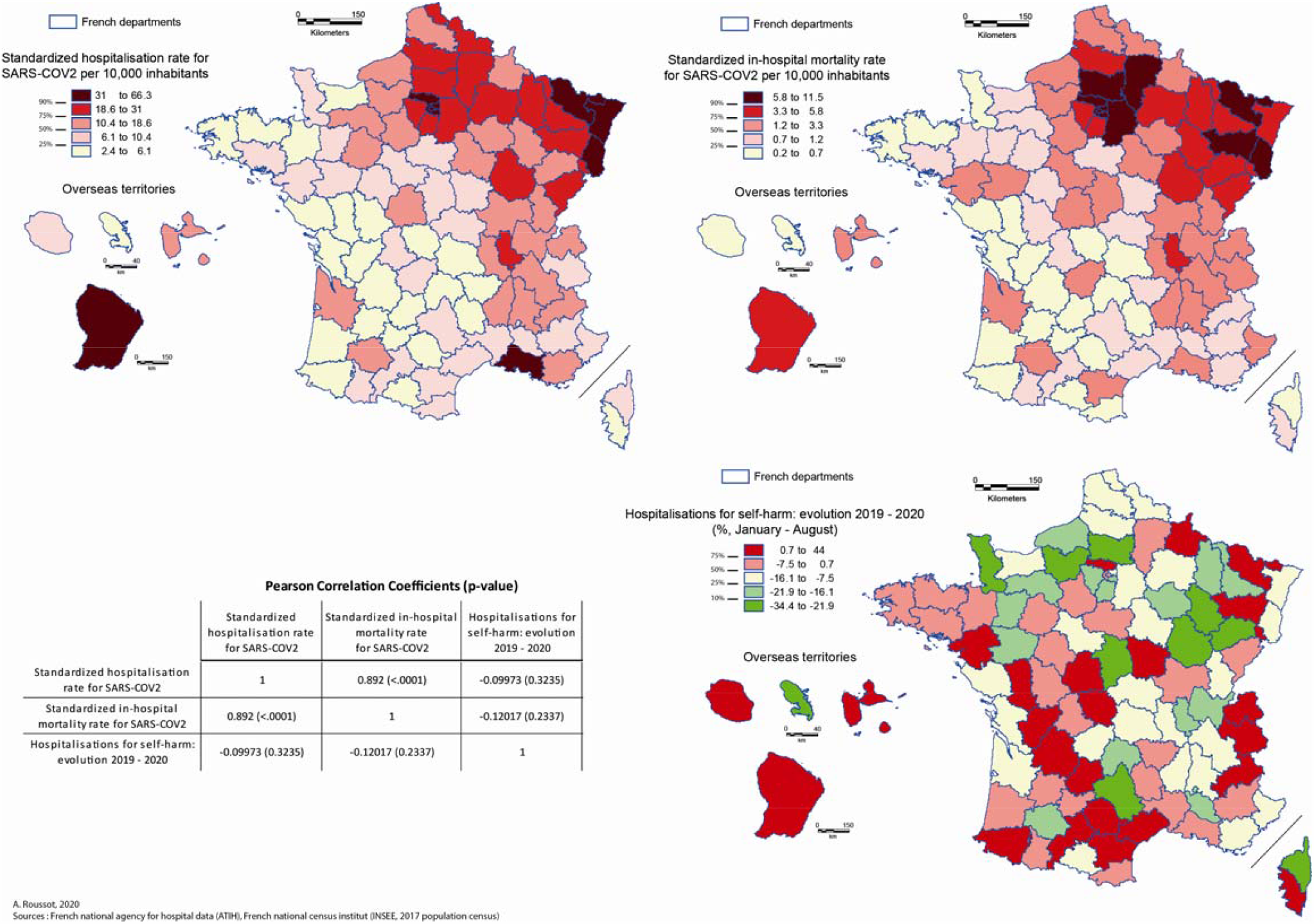
Maps of rates of hospitalization and mortality due to Covid-19, and change in the number of hospitalizations for self-harm in France from 2019 to 2020 (January to August only) in each French administrative department.

## DISCUSSION

While the Covid-19 pandemic and the quarantine period were generally associated with increased signs of anxiety and depression across countries including France, our data suggests that it did not translate into increased hospitalizations for self-harm overall. On the contrary, we observed an 8.5% decrease in the number of hospitalizations between 2019 and 2020. A decrease was also found when comparing data to 2018 and 2017. Moreover, it precisely started the first week of the confinement and persisted after its end and until the end of the observation period in August. One hypothesis may be that the lockdown of several individuals at home may have prevented many acts to occur. Also, some previous reports suggest that self-harm decrease in times of national tragedies, the so-called “pulling-together effect” ^12, 25^. It is, however, important to keep in mind that during this period of more limited access to care and emergency departments ^26^, many people who self-harmed may have stayed at home. One study in France before the Covid-19 pandemic showed that up to 40% report a history of self-harm without subsequent presentation to the hospital ^27^.

Two important exceptions to this decrease in self-harm hospitalizations have to be highlighted. First, an increase in hospitalizations was observed in individuals aged 75 and older (and even 65+), both in men and women. This age group has been the most impacted by the Covid-19 pandemic in terms of mortality and hospitalizations. It may also have particularly suffered from restrictions in travels and visits, favoring social isolation. Increased vigilance, care and social support to the elderly are therefore necessary.

Moreover, we observed an increase in more severe suicidal acts, including more hospitalizations in intensive care for self-harm in 2020 than 2019, more self-harm using firearm, drowning or jumping from height, and more deaths at hospitals following self-harm. The decrease in overall self-harm hospitalization may therefore have mainly concerned less lethal acts while the number of severe gestures with high lethality rose. This is a very concerning observation that may be confirmed in some months when data on suicide completion will be available. Until then, suicide prevention strategies should continue to be implemented and run such as “VigilanS”, a national surveillance program for people who self-harmed ^28^, and others.

It is interesting to note a total decoupling across administrative departments between the level of severe cases of Covid-19 contamination ^29^, and the impact on self-harm hospitalizations, suggesting that the reality of the local risk of the pandemic had no or limited effects on self-harm occurrence overall. Indeed, during the early months of the pandemic in France, some regions showed a high level of contamination (roughly, the North-Eastern part of mainland France, and Guyana) with media reports of overwhelmed hospitals and intensive care units while other regions, that were equally confined, had very low levels. Self-harm result from the complex interaction of many factors including life events, mental illness and a personal history and vulnerability ^30^.

Results presented here refer to the early period of the pandemic including the first confinement. Observations may be different during the following months. It is now obvious that the pandemic will likely last several more months (which led to a new confinement at the end of 2020), and many people find themselves in difficult financial situations with limited hopes that the national economy will improve soon. Students have also been particularly affected by the situation in relation to disturbed conditions of teaching, limited access to some jobs (e.g. in restaurants and bars), and reduced social life. Finally, the brain and psychological consequences of severe Covid-19 infection are still unknown. A 12 year follow-up of a cohort of individuals infected by the severe adult respiratory syndrome (SARS) outbreak in 2003 previously showed increased rates of mental disorders and suicide ^31^. Regarding Covid-19, early investigations report up signs of psychological distress in up to 40% of survivors ^32^ raising concerns about the long-term effects of the infection. Suicide prevention should therefore remain a priority, both in the short- and long-terms.

## Data Availability

National administrative data were used and are available upon request to the administration.

## Notes

### Competing Interest Statement

The authors have declared no competing interest.

### Funding Statement

None

### Author Declarations

approved by the National Committee for data protection (registration number 2204633)

